# Heparin therapy improving hypoxia in COVID-19 patients - a case series

**DOI:** 10.1101/2020.04.15.20067017

**Authors:** Elnara Marcia Negri, Bruna Mamprim Piloto, Luciana Kato Morinaga, Carlos Viana Poyares Jardim, Shari Anne El-Dash Lamy, Marcelo Alves Ferreira, Elbio Antonio D’Amico, Daniel Deheinzelin

## Abstract

**INTRODUCTION:** Elevated D-dimer is a predictor of severity and mortality in COVID-19 patients and heparin use during in-hospital stay has been associated with decreased mortality. COVID-19 patient autopsies have revealed thrombi in the microvasculature, suggesting intravascular coagulation as a prominent feature of organ failure in these patients. Interestingly, in COVID-19, pulmonary compliance is preserved despite severe hypoxemia corroborating the hypothesis that perfusion mismatch may play a significant role in the development of respiratory failure.

**METHODS:** We describe a series of 27 consecutive COVID-19 patients admitted to Sirio-Libanes Hospital in São Paulo-Brazil and treated with heparin in therapeutic doses tailored to clinical severity.

**RESULTS AND DISCUSSION:** PaO2/FiO2 ratio increased significantly over the 72 hours following the start of anticoagulation, from 254(±90) to 325(±80), p=0.013, and 81% of the patients were discharged home within a mean time of 11.4 (±7.9) days. Most mechanically ventilated patients (67%) were extubated within 12.5(± 5.7) days. There were no bleeding complications or fatal events.

Even though this uncontrolled case series does not offer absolute proof of DIC as the underlying mechanism of respiratory failure in COVID-19, patient’s positive response to tailored dose heparinization contributes to the understanding of the pathophysiological mechanism of the disease and provides valuable information for the treatment of these very sick patients while we await the results of further prospective controlled studies.

## INTRODUCTION

Since the beginning of the COVID-19 pandemic, disease severity has been linked to markers of coagulation disturbances such as prothrombin time prolongation, elevated fibrin degradation products, reduced platelet count, and specially to elevated D dimer ^1-7^. Higher levels of D dimer and the presence of other coagulation disturbances have been independently associated with development of respiratory failure and death in patients with COVID-19^8^. The use of heparin, particularly in those patients with more pronounced elevations of D dimer and in those with elevated sepsis induced coagulopathy (SIC) score, has been associated with a better prognosis. Diabetic patients, whose levels of D dimer are greater than those of non-diabetic patients, have also been shown to have a worse prognosis regarding COVID-19^4 8^. Moreover, D dimer levels have helped differentiate severe COVID-19 associated pneumonia from that caused by other viruses^10^.

Over the past 2 months it has been consistently shown that SARS-Cov-2 causes a cytokine storm that ultimately leads to the activation of the coagulation cascade, causing thrombotic phenomena^4 8 11^. Similarly to what happens in severe sepsis, the widespread deposition of intravascular clots compromises adequate blood supply, contributing to organ failure^12^.

Disseminated intravascular coagulation (DIC) secondary to severe infection is classically associated with gram-positive or gram-negative bacteria, malaria and haemorrhagic fevers, but other viruses, such as dengue (an hemorrhagic virus), SARS-CoV and MERS-CoV, can also be responsible for systemic activation of intravascular coagulation^13 14^.

Furthermore, in contrast to the characteristic stiffening of the lung usually seen in acute respiratory distress syndrome (ARDS), in COVID19 patients the severe hypoxemia observed is accompanied by near normal pulmonary compliance, especially in early stages^15^. Autopsy findings from COVID-19 patients show microthrombi in the pulmonary microvasculature^16-18^ suggesting that ventilation-perfusion mismatch due to capillary obstruction could be a pivotal feature in the refractory hypoxemia presented by these patients. The anatomical distribution of this peripheral vascular bed mirrors the predominantly distal and patchy distribution of the radiological infiltrates^19^.

In one of our first COVID-19 patients we noticed a concomitance of peripheral ischaemia (acro-ischemia) with the onset of respiratory distress, an observation that led us to consider the hypothesis that the normal compliance respiratory failure might actually be due to extensive pulmonary capillary obstruction, and that systemic disseminated intravascular coagulation might be playing a significant role in hypoxemia and outcome of COVID-19 patients.

The treatment of DIC consists in slowing down the coagulation cascade by using low doses of anticoagulation, alongside vigorous specific treatment of the underlying disorder. We therefore considered adding early heparin therapy to our standard care^2^. The present study is a description of the outcome of the first 27 COVID-19 patients we treated as DIC in the course of the disease.

## METHODS

This study is a case series of 27 consecutive Covid-19 patients seen by our team in Sirio-Libanês Hospital – São Paulo, Brazil, between March 21^st^ and April 12^th^, 2020. The study is registered under number 1678 at the Sirio-Libanês Hospital and informed consent was waived.

All patients with COVID-19 admitted by our team received enoxaparin 0,5mg/kg SC every 24 hours. Patients with a creatinine clearance under 30 mL/min received subcutaneous unfractionated heparin at a dose of 5,000 units every 8 or 6 hours. If an abrupt decrease in oxygenation or an increase in D Dimer levels was observed, enoxaparin dose was raised to 0,5 mg/kg SC every 12 hours and, in the event of thrombotic phenomena or worsening hypoxia, the dose was further increased to 1 mg/kg SC every 12 hours. Patients with a BMI (body mass index) of 35 or higher were also considered for the higher dose regimen. Patients in shock were treated from the beginning with intravenous heparin, targeting an APTT ratio around 1.5 to 2.0 times the normal range. If patients presented any acute thrombotic event, heparin dosing was increased to obtain an APTT approximately 2.0 to 2.5 times the normal range.

All patients received a 10-day course of azithromycin (500mg on day 1, then 250mg daily)^20^. Methylprednisolone 40mg daily was initiated if a worsening in the radiological pattern accompanied by an increase in serum LDH levels was observed. If the patient presented subsequent rise in C-reactive protein, we actively searched for secondary infection and promptly initiated antibiotics.

## RESULTS

We followed a total of 27 hospitalized patients with a diagnosis of COVID-19, all confirmed by PCR. Seventy percent were male, their mean age was 56 ± 17 years, mean BMI was 28.8 ± 6 kg/m^2^, and comorbidities were present in 56% of them (table 1). The mean WHO classification score^3-7^ at admittance was 4.0 ± 1,2 (Figure 1). Entry CT scans showed radiologic infiltrates compromising up to 25% of lung area in 22% of patients, 25-50% of lung area in 48% of patients, and 30% of patients presented infiltrates in over half of lung parenchyma. Symptoms started at an average of 9.6 ± 4.0 days prior to hospitalization, and the anticoagulation protocol was initiated at an average of 3,4 ± 4,0 days after admission. Nineteen patients received methylprednisolone in the course of the disease. The average level of D Dimer during the follow up period was 1,637± 1,967 ng/mL FEU, with a peak value of 3,544 ± 5,914 ng/mL FEU. Only 5 patients (19%) never had a D dimer level above 500 ng/mL FEU. Six patients received only the prophylactic dosage of heparin or enoxaparin; three patients started already with enoxaparin 0,5 mg/Kg twice and were kept on this dosage and in 18 patients’ dosages were escalated to either full EV heparin or enoxaparin 1mg/kg twice a day.

**Table 1.** Comorbidities incidences

| **Comorbidities - n(%)** |  |
| --- | --- |
| Total=15 patients (56%) |  |
| Diabetes | 3 (11%) |
| Hypertension | 7 (26%) |
| Heart disease | 3 (11%) |
| Previous lung disease | 2 (7%) |
| Cancer | 1 (4%) |
| Other | 7 (26%) |

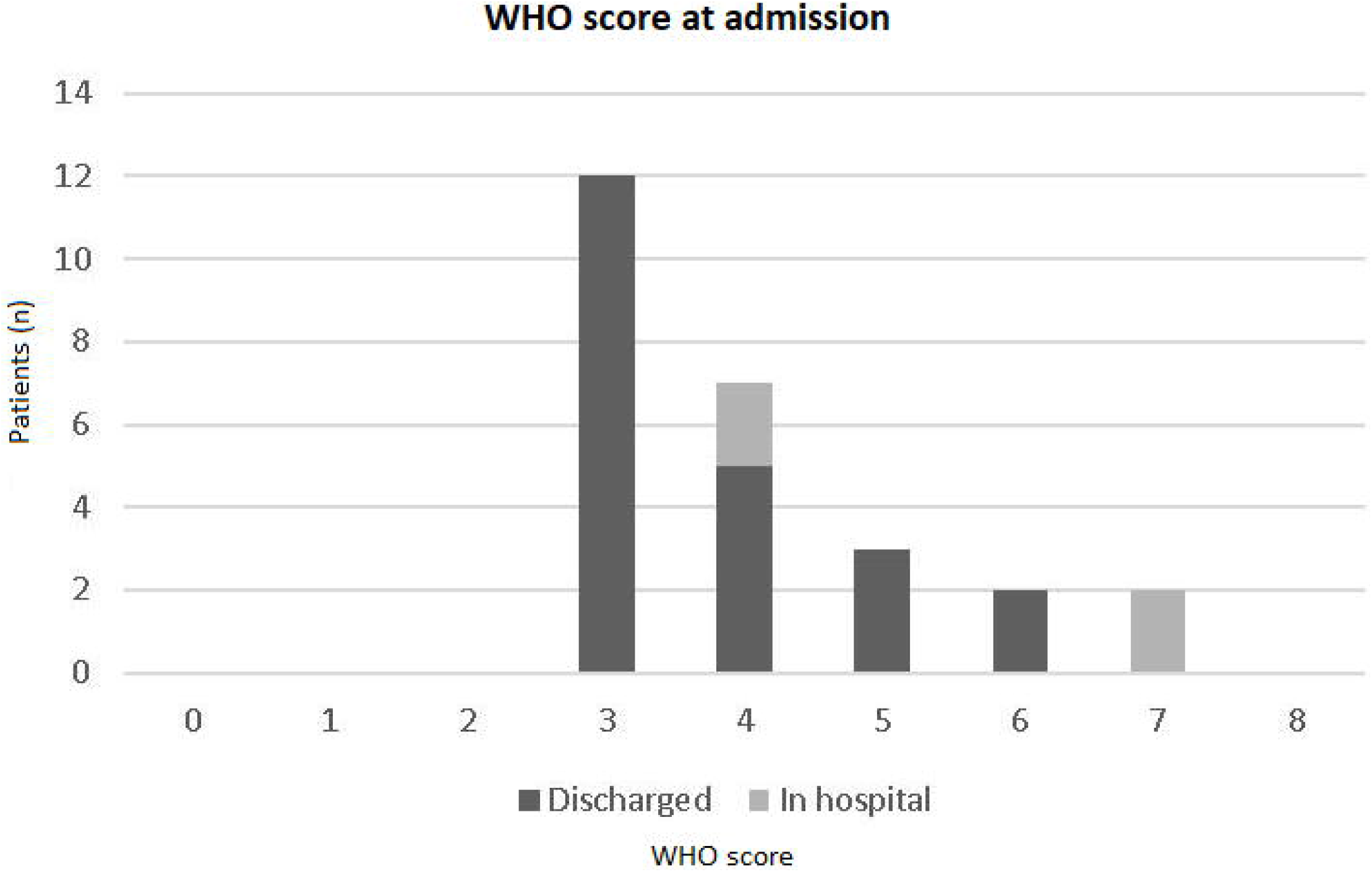

Of the 27 consecutive patients, 22 (81%) were discharged from hospital after an average of 11.4 (± 7.9) days. One patient was transferred to another hospital on the 4th day and lost follow-up. Nine patients (33%) were admitted to ICU, 6 (66%) of which have already been discharged to the ward after an average of 13.2(±6.7) days. Eight patients (30%) required intubation, and 5 patients have already been successfully weaned after an average 12.5 (± 5.7) days of mechanical ventilation. Of the patients still under mechanical ventilation, 2 required a tracheostomy.

Interestingly enough, rotational thromboelastometry (ROTEM) performed in four patients, showed an increase in α-angle, amplitude 10 minutes after clotting time (A10) and maximum clot firmness (MCF) pointing to a persistent hypercoagulability state, despite their ongoing heparin use.

Figure 2 depicts the gradual improvement in PaO_2_/FiO_2_ ratio along the first 72 hours in relation to pre-anticoagulation values. Analysis was conducted for the whole series (A) and considering only patients with moderate to severe disease (B) according to the WHO score (p<0,02 for both groups). For non-mechanically ventilated patients PaO_2_/FiO_2_ ratio was calculated according to mask or nasal catheter oxygen flow and oxymetry^21^.

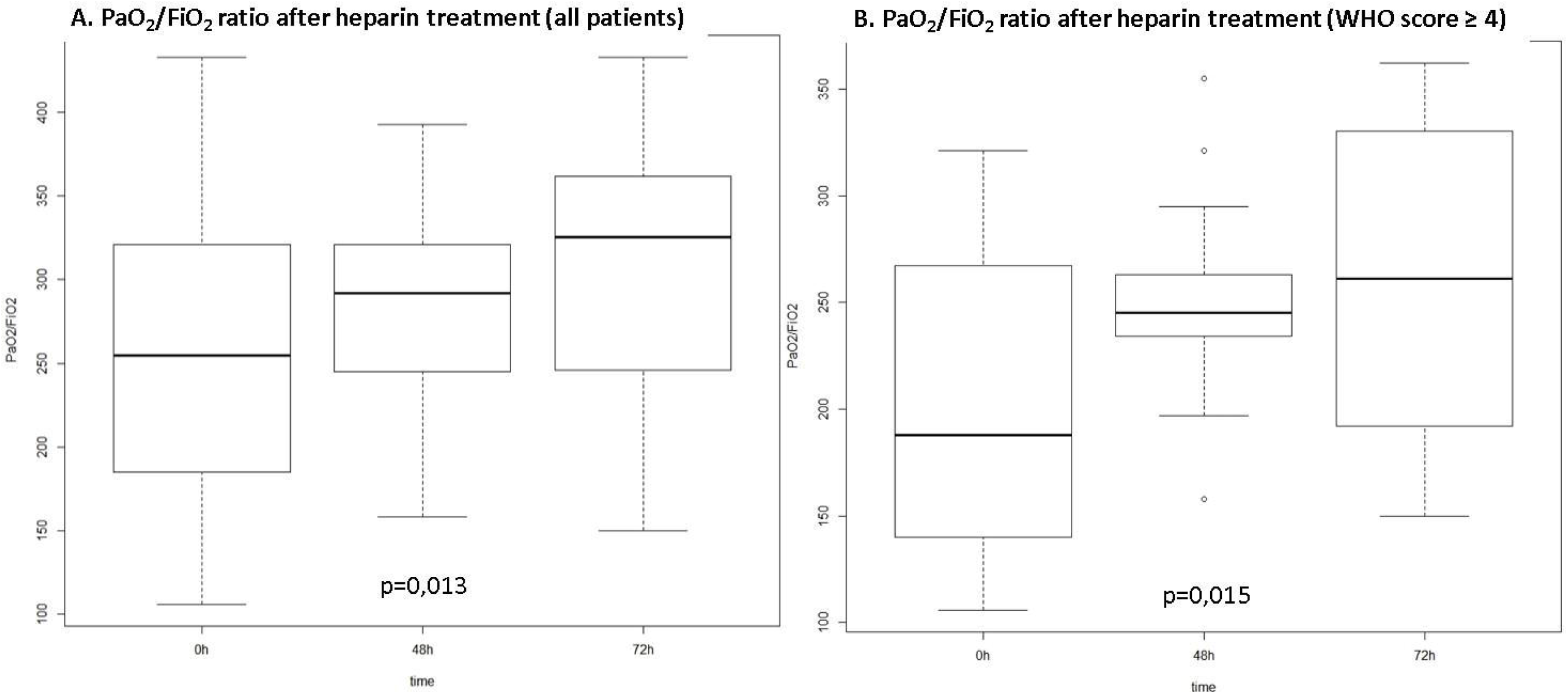

We observed no deaths or haemorrhagic complications due to anticoagulation during the study period.

## DISCUSSION

Our results suggest the important role of disseminated intravascular coagulation as one of the main mechanisms of organ failure in COIVID-19 and the potential response to early anticoagulation therapy. The significant improvement observed in oxygen exchange and clinical symptoms observed in these COVID19 patients in response to the anticoagulation points to a potential role for systematic use of heparin in the treatment of such patients. The high incidence of thrombotic events that has been reported in COVID-19 patients^22^, as well as the fact that similar observations were reported in the other recent coronavirus outbreaks^13 14^, further corroborate with this line of reasoning. This is not surprising, as severe cases of COVID19 meet the laboratory criteria of DIC^4 8^ of thrombotic pattern, in which fibrinogen does not drop and prothrombotic phenomena override the haemorrhagic ones^23^. The histological findings of thrombi in the microvasculature documented in the autopsies of COVID-19 patients^17^, further support the belief that in COVID-19 we are facing a thrombotic organ-failure type of DIC that should be treated with heparin^23 24^. This might explain the previous findings of an association between heparin use during hospital stay and a reduced mortality^4^.

Thromboelastography showing a pattern of hypercoagulability despite the use of heparin during the course of viral diseases has been previously reported^25^. In fact, many viruses known to induce a state of hypercoagulability^26^ have a similar pattern of disease, including the timeframe of clinical manifestations^27^, suggesting a common pattern of response.

Although the principle that early anticoagulation therapy for sepsis probably causes uncontrolled immunothrombosis and pathological systemic DIC, since the presence of neutrophil extracellular traps (NETs) and hypercoagulation in DIC localize the infection, this is well described for bacteria but not for virus, where virus-induced NETs could be a pathogenic mediator^28^.

The PaO_2_/FiO_2_ ratio improvement observed in our patients after starting heparin is in agreement with the idea of a significant perfusion component explaining the mechanism of respiratory failure with the distinct pattern of marked hypoxia and preserved lung compliance that characterizes severe COVID19 patients. It has been argued that this could be secondary to the loss of lung perfusion regulation and hypoxic vasoconstriction^16^, but the clinical response to heparin rather suggests hypoxia due to extensive clogging of pulmonary microcirculation. Interestingly the use of tissue Plasminogen Activator (tPA) has been shown to promote a non-sustained elevation of PaO2/FiO2 ratio^29^. In our opinion, given the marked hypercoagulability seen in these patients - and again in accordance with the autopsy findings - judicious tailoring of heparin doses is needed to prevent capillary reocclusion while avoiding the risks of bleeding complications.

The fact that this is a retrospective study without a control arm does not yet allow us to definitively conclude that heparin in tailored doses should be systematically employed in all COVID19 patients. Nonetheless, our findings in this early group of patients certainly provide food for thought and perhaps a rationale to justify using a readily available and well-known drug such as heparin to ameliorate the dim prognosis of such sick patients while we await the more solid data on this subject.

## Data Availability

Data will be provided if requested. It contais confidential medical records.

## ACNOWLEDGMENTS

The authors would like to thank the Teaching Research Institute (IEP), Sirio-Libanes Hospital, Sao Paulo, Brazil. We also would like to acknowledge the hospital staff and their instrumental role in caring for all patients.

## AUTHOR’S CONTRIBUTIONS

EMN designed and provided the original idea for the stydy; BMP and LTKM collected the data and provided the rationale for all analyses; CVPJ, DD, EB revised the article and contributed for the discussion. SAEDL and MAF conducted the statistical analyses. All authors reviewed the final manuscript.

